# On the impact of early non-pharmaceutical interventions as containment strategies against the COVID-19 pandemic

**DOI:** 10.1101/2020.05.05.20092304

**Authors:** Andrés Hernández, Esteban Correa-Agudelo, Hana Kim, Adam J. Branscum, F. DeWolfe Miller, Neil MacKinnon, Diego F. Cuadros

## Abstract

**Background:** The novel coronavirus SARS-CoV-2 (COVID-19) emerged in December 2019 in Wuhan, China and has spread since then to around 210 countries and territories by April 2020. Consequently, countries have adopted physical distance measures in an attempt to mitigate the uncontrolled spread of the virus. A critical question for policymakers to inform evidence-based practice is if and how physical distance measures slowed the propagation of COVID-19 in the early phase of the pandemic.

**Methods:** This study aims to quantify the effects of physical distance mitigation measures on the propagation of the COVID-19 pandemic. Data from John Hopkins University on confirmed cases and testing data from the Our World in Data were used in an interrupted time series analysis to estimate the effects of physical distance measures on the growth rates of the pandemic in 12 countries of Asia, Africa, and Europe.

**Findings:** We found that physical distance measures produced a significant decrease in the growth rates of the COVID-19 pandemic in five countries (Austria, Belgium, Italy, Malaysia, and South Korea). The test-positivity rate was significant in understanding the slowing growth rate of COVID-19 cases caused by the mitigation measures, as it provides important context that is missing from analysis based only on confirmed case data.

**Interpretation:** Physical distance interventions effectively slowed the progression of the COVID-19 pandemic. The results of this study could inform infectious disease mitigation policies based on physical distance measures by quantifying the differential health outcomes of a pandemic with and without physical distance interventions.

**RESEARCH IN CONTEXT:** *Evidence before this study:* The SARS-CoV-2 is a new virus identified in December 2019 in the province of Wuhan, China and as never before, a remarkable number of studies and reports have been released since the start of the pandemic. Several studies have used confirmed COVID-19 cases to estimate the growth rate of the pandemic. However, many studies have discussed limitations of including only confirmed cases attributable to the lack of information about testing protocols and testing rates among different countries. Finally, some researchers proposed the analysis of reported deaths by COVID-19 as a potential solution. However, this metric results in biased estimates because deaths by COVID-19 are known to be underreported.

*Added value of this study:* We designed and implemented analytic methods based on our previous research applied to different infectious disease epidemics, to add evidence related to the impact of non-pharmaceutical containment strategies on the temporal progression of the COVID-19 pandemic. Specifically, this study adds quantitative evidence about the effects of physical distance measures on limiting the propagation of COVID-19 pandemics in different countries. Additionally, we included testing data in the analysis to assess intra- and inter-country variation in testing growth rates. We hypothesized that the test-positivity rate is an approximation to the incidence of the COVID-19 pandemics in countries with high testing rates. Additionally, we hypothesize that a significant decrease in the pandemic over time could be identified by a significant decrease in the confirmed cases along with a significant decrease in the test-positivity rate. Our results quantified the potential effects of physical distance interventions on the COVID-19 pandemic progression under different levels of testing and enforcement of mitigation policies.

*Implications of all the available evidence:* Our analysis could lead to better approaches for estimating the effects of physical distance measures on the time course of infectious diseases. In addition, our analysis highlights the potential bias of estimated COVID-19 growth rates based only on confirmed cases. The results from our study could inform strategies for mitigating the COVID-19 or other future pandemics, especially in countries in an earlier stage of a pandemic.

## INTRODUCTION

Early data on the novel Coronavirus disease (COVID-19) from Europe and Asia show the exceptionally high contagious rates of the pandemic^1^. In late March 2020, the United States had the fastest growing curve in terms of incident COVID-19 infections across developed countries. Under this scenario, quick remedial measures taken by several countries (e.g., China, Italy, and Spain) included restrictions on physical interaction and public events followed by full physical isolation lockdowns as non-pharmaceutical interventions (NPI)^2–4^. Conversely, other countries, including the United States, adopted softer geographically dependent measures in terms of physical interaction and isolation^5^.

With COVID-19 rapidly spreading^6^ and with treatment and a vaccine still under early stages of development, a critical question for policymakers is how NPIs can reduce the growing confirmed number of incident cases and the test-positivity rate (TPR) (i.e., the proportion of positive tests among those people who were tested for COVID-19) in the early and mature phases of the pandemic^2,7,8^. Such information might not only justify physical isolation policies, but also provide time to adapt or redistribute health care personnel, technology, and services where they are needed the most. These analyses would also provide crucial benchmarks for measuring the impact of potential relaxation of the physical distancing interventions in the coming months of the pandemic. In this study, we evaluated the impact of NPI protocols, including lockdown and physical distancing efforts, to counteract the initial intensity of COVID-19 outbreaks in several countries. We also conducted a counterfactual analysis to estimate the number of COVID-19 cases that would occur if such NPI protocols were not implemented in 12 developed countries in Europe, Asia, and Africa.

## METHODS

### Data sources

We obtained the number of daily confirmed cases of COVID-19 by country from datasets provided by the Johns Hopkins University Center for Systems Science and Engineering^9^ (JHU CSSE). The number of confirmed cases reported at the country level was obtained from twitter feeds, online news services, and direct communication. The JHU CSSE team obtains COVID-19 case confirmation from regional and local health departments, the European Center for Disease Prevention and Control, the World Health Organization (WHO COVID-19 Report), as well as from city and state health authorities.

The test-positivity rate for COVID-19 was calculated by country using testing count data from the Our World in Data Project^1^. The TPR is defined as the number of laboratory-confirmed positive tests per 100 suspected cases examined, and is a metric used by infectious diseases surveillance programs as one of several indicators of temporal trends of epidemics^10^. The dataset is available in the GitHub repository for COVID-19 testing conducted in 40 countries. Details on COVID-19 case counts and testing counts are included in the documentation of the repository and correspond to official government policy for each country.

### Data analysis

Temporal trends of the number of accumulated confirmed COVID-19 cases until April 10^th^, 2020 were identified using a Bayesian interrupted time series analysis implemented in the CausalIimpact^11^ package in R version 3.5.2. This analysis quantifies significant changing trends in COVID-19 rates (over a 12-day period) from the rates expected in the absence of any enforced interventions, namely physical distancing, stay home orders and lockdowns. Using this method, we simulated changes in the trajectory of the COVID-19 pandemic from the first to the last dates in which testing data were available by country. Accumulated testing data were included only for the countries with enough daily test information to allow reasonable interpolation. As a result, 12 countries from Europe, Asia, and Africa were included in our analysis: Austria, Belgium, Estonia, Finland, France, Iceland, Italy, Japan, Malaysia, South Africa, South Korea and the United Kingdom. We interpolated testing data for the dates with missing information using a Kalman filter approximation^12^. The number of confirmed COVID-19 cases divided by the corresponding interpolated testing count gave the TPR by day, which were resampled every two days to avoid high variability. Finally, the resulting resampled time series for each country was linearized, and growth rates were obtained using the natural logarithm and the discrete first derivative.

Accumulated cases and testing data were analyzed to account for the time lag between testing and confirmation of COVID-19 cases. Our analysis assumed that both the number of cases and the number of tests were exponential functions of time because the COVID-19 epidemic was still under the exponential growth phase at the time of analysis. The counterfactual trend analysis quantifies the differences between the observed COVID-19 growth rates and the (unobservable) growth rates that would have occurred under the absence of physical distancing measures. We conducted the analysis for each day in which data were available to identify dates where differences were statistically significant after NPI measures were implemented. The analysis was conducted independently for confirmed cases and TPR in order to assess the impact of different levels of testing in the reporting of the COVID-19 pandemic. Our Bayesian state-space time series analysis constructed counterfactual trends by using the rates before NPI measures were implemented to estimate the alternative outcome without the effect of the intervention. This methodology quantifies the deviation of the time series during the post intervention period from its baseline. This model is valid for short-term predictions, and used default prior distributions in the causal impact model^11^. COVID-19 cases under the counterfactual model were computed from their posterior predictive distributions, and the difference in growth rates between the observed values and the counterfactual estimates were used to identify significant effects of physical distance measures to counteract COVID-19 transmission. The benchmark for significant changes was set to 0.5 standard deviations from the pre-intervention values in order to increase the robustness of results. Data from a period of six days before to six days after the first significant change in growth rates after implementation of NPI intervention were used to estimate the observed and counterfactual percent differences in the growth rates of COVID-19 cases.

Finally, we created tabular comparisons of the estimated and actual growth rates (for confirmed cases and testing rates), dates of first case confirmed and physical distance measures^2^, and level of testing per one thousand people by country. Statistical analyses were conducted using R version 3.5.2^13^ (R Project for Statistical Computing), statistical significance was set at P< 0.05 where P is the Bayesian one-sided tail-area posterior probability, and graphs of times series were created using the R package ggplot2^14^

## RESULTS

### Descriptive statistics of COVID-19 in the selected countries

Descriptive statistics for the COVID-19 pandemic in the 12 countries selected for our analysis are presented in Table 1. These countries reported a total of 385,261 COVID-19 confirmed cases and 2,420,053 tests by April 10^th^, 2020. The total number of reported COVID-19 deaths was 41,169, which was 10.68% of the confirmed cases. Average TPR at the last available date was 12.30% (standard deviation [SD] 10.97%) for all countries included in our analysis.

**Table 1.**
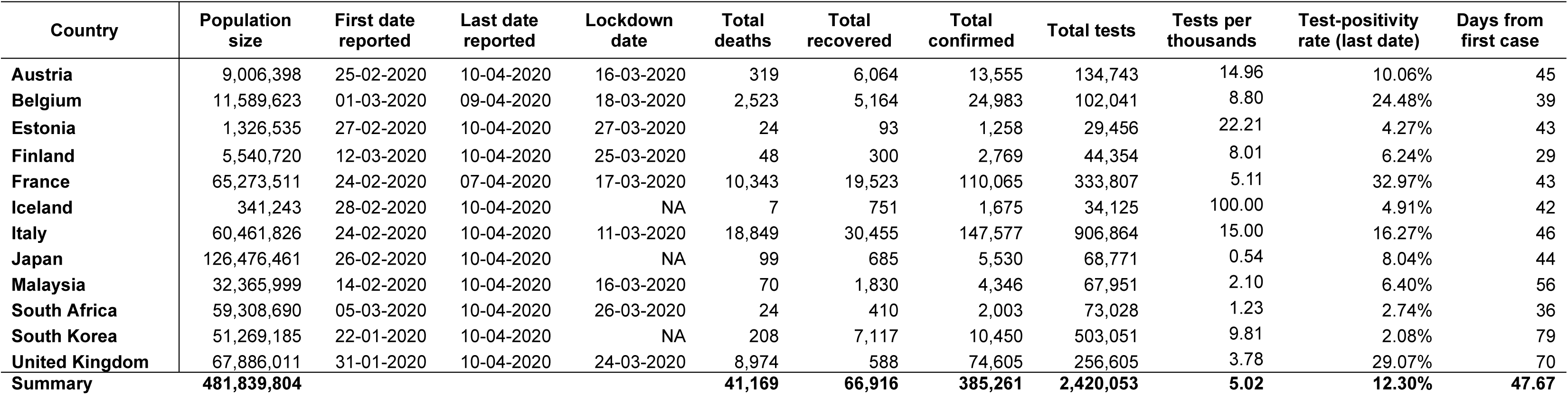
Descriptive statistics of the COVID-19 pandemic by country

The total COVID-19 testing per 1,000 individuals was 5.02 (1,000 * 2,420,053 / 481,839,804), and the country with highest testing per 1,000 inhabitants was Iceland (100.00), followed distantly by Estonia (22.21) and Italy (15.00). The highest TPR occurred in France (32.97%), the United Kingdom (29.07%), and Belgium (24.48%). Countries with the lowest TPR were South Korea (2.08%), South Africa (2.74%), and Estonia (4.27%).

### Time trend analysis of COVID-19 cases

Table 2 presents results from time trend analysis of confirmed cases and the TPR on the day the first significant effect was detected after NPI measures were enforced. Physical distancing interventions were applied on average 18.92 days (SD 14.06 days) after the first COVID-19 case was reported. On average, a significant decrease in growth rates of the pandemic was observed 12.33 days after implementation of NPI measures (SD 5.61 days).

**Table 2.**
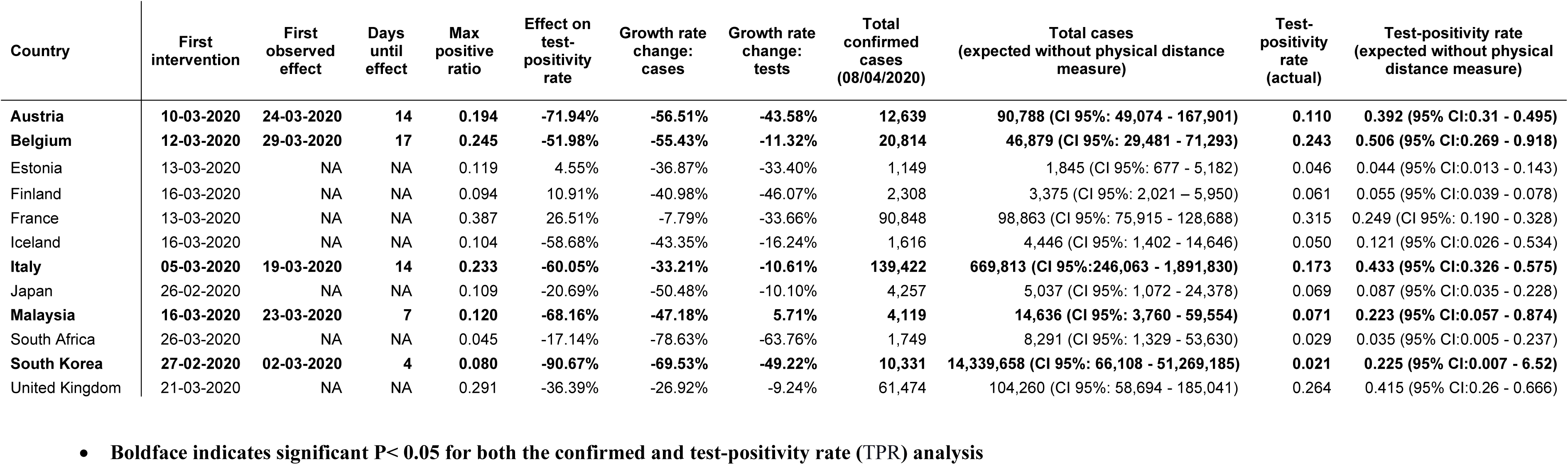
Estimated effects of physical distance measures on COVID-19.

Significantly decreasing growth rates of COVID-19 after mandating physical distancing were observed in five countries (Austria, Belgium, Italy, Malaysia, and South Korea), with an average reduction of 52.37% (SD 13.37%) in the growth rates of confirmed cases. For these five countries, there was a decrease of 23.22 (95% confidence interval [CI] 17.87 - 28.57) cases per 100 tests, after implementation of NPI measures. Conversely, seven countries (Estonia, Finland, France, Iceland, Japan, South Africa, and the United Kingdom) did not have significant changes in the growth rate of COVID-19 after implementation of NPI measures (controlling for temporal changes in testing rates).

The estimated temporal trends of confirmed COVID-19 cases and the TPR in the absence of physical distancing, compared to the observed values, are presented in Figure 1. The results in Figure 1 and Table 2 identify the countries with significant declines in growth rates of the pandemic attributable to NPI interventions (see graphs for Austria, Belgium, Italy, Malaysia and South Korea in Figure 1). On average, the percent decrease between the expected TPR in the absence of NPI intervention and the actual ratio in the countries with a significant decline in growth rate was 68.56% (SD 14.56%), compared to a much lower reduction for the countries without significant effects 12.99% (SD 29.30%).

**Figure 1.**
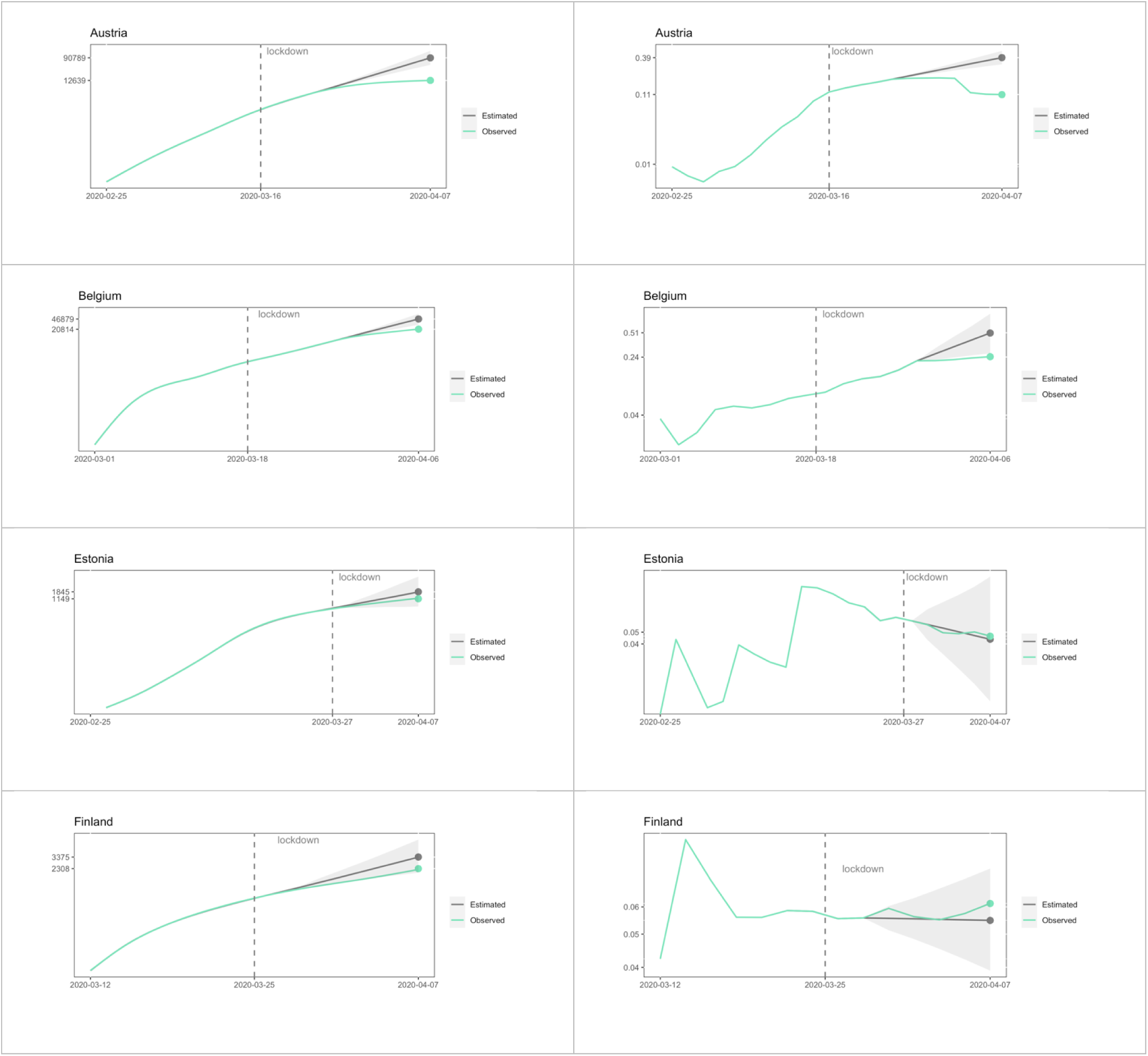
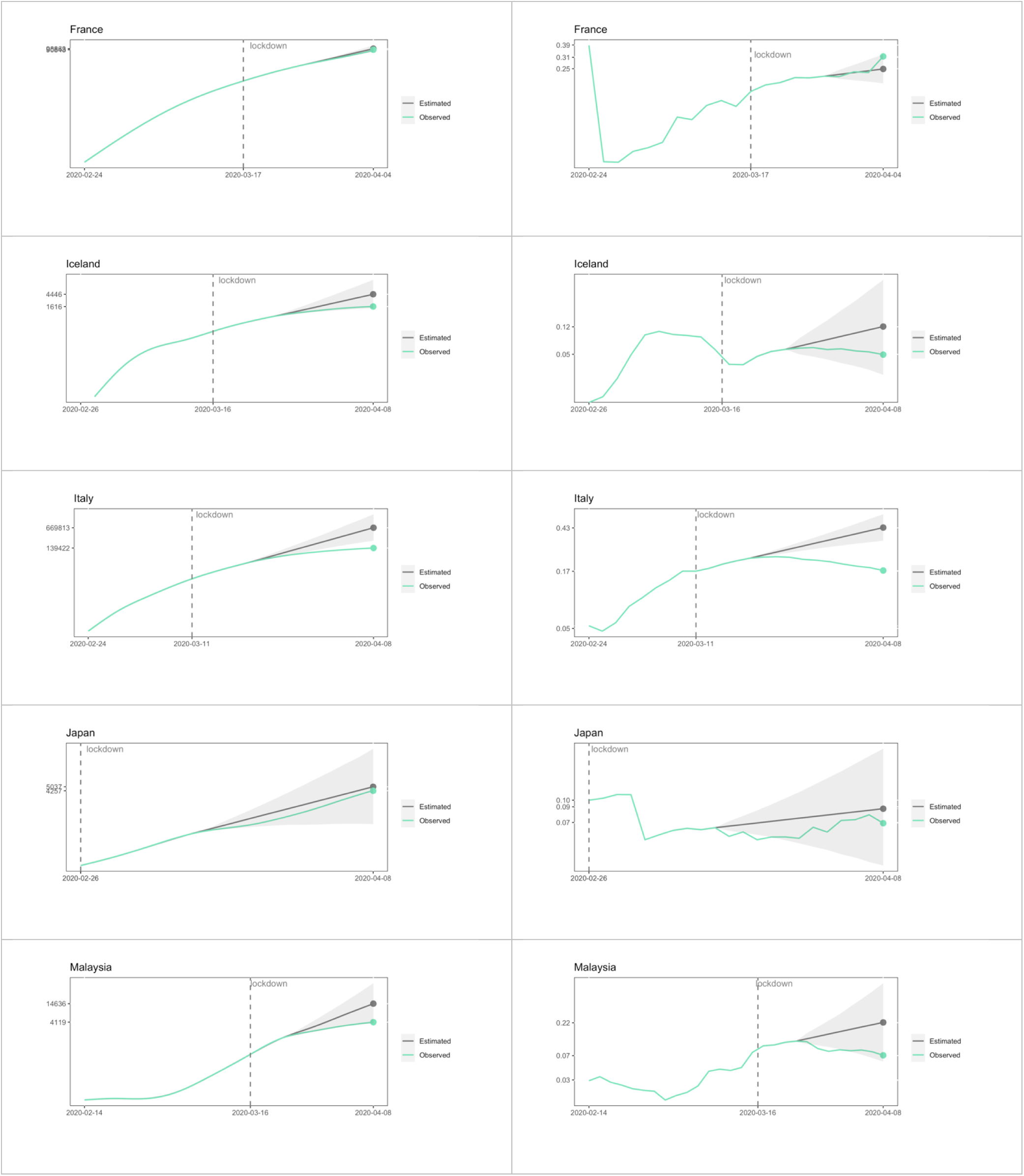
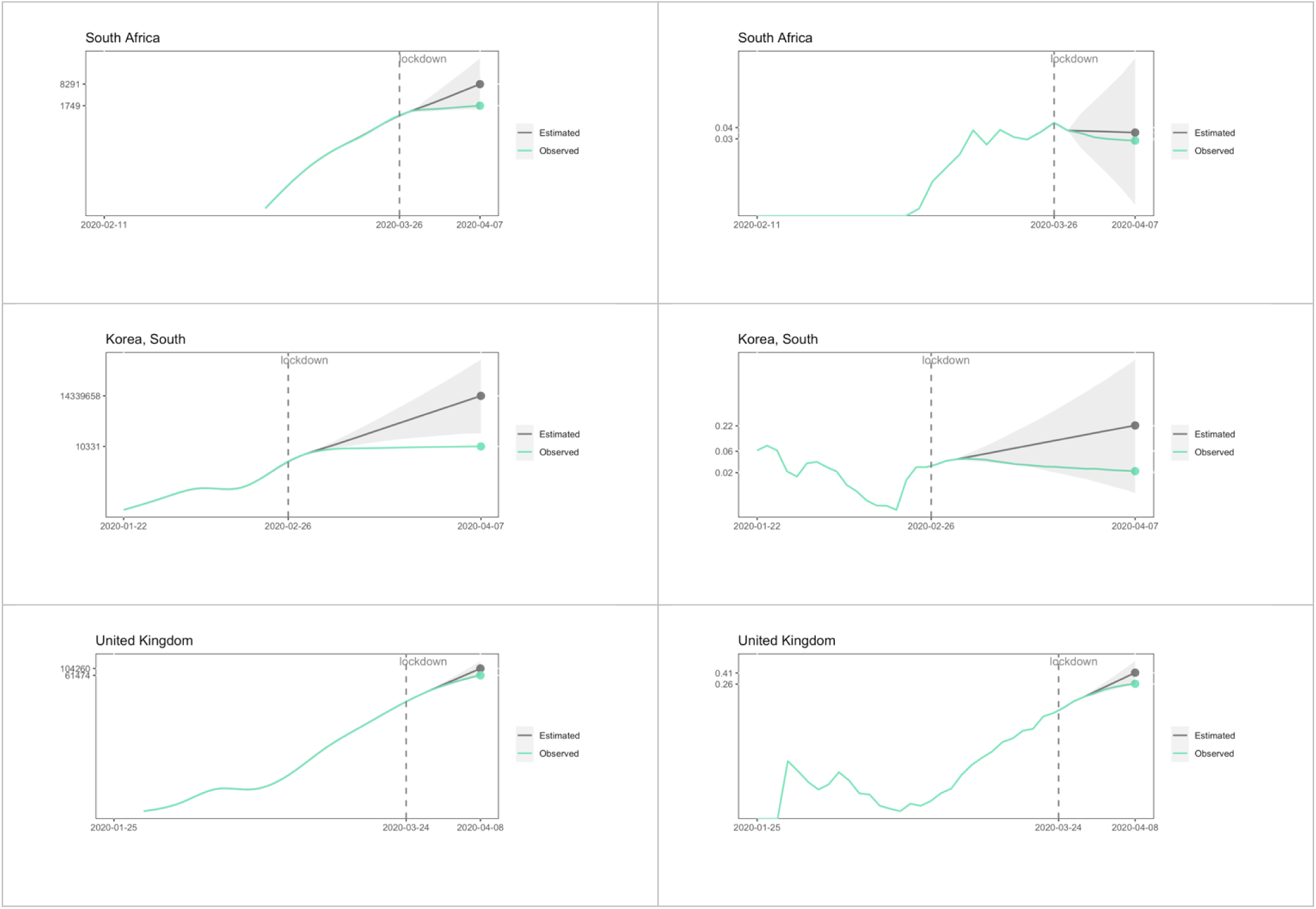
Observed COVID-19 confirmed cases (left column) and test-positivity rate (TPR, right column), and estimated values in the absence of physical distancing measures by country.

## DISCUSSION

Using available data for the COVID-19 pandemic on confirmed cases and testing counts in 12 countries in Europe, Asia, and Africa, we estimated the potential effects of NPI mitigation policies such as physical distancing and lockdowns on COVID-19 growth rates by country after controlling for temporal changes in testing rates. Our results suggest that these NPI measures contributed to the reduction of cases and TPR in several countries, depending on the NPI duration and the intensity of enforcement of NPI policies. Moreover, time trend analysis of TPR data identified countries where the observed reduction in the speed of the spread of COVID-19 might be a result of changes in the testing rate over time, and not the result of the NPI measures implemented. Our results support the analysis of TPR data in combination with time series data on confirmed cases in a country instead of the analysis of confirmed cases alone.

We estimate that NPI measures such as physical distancing and lockdowns led to an average reduction of 39% to 65% in the growth rate of confirmed COVID-19 cases. This effect along with a reduction of 23.22 cases per 100 tests suggests that these NPI measures are contributing to flattening the curve of the COVID-19 distribution. The results of the application of these NPI measures are evident in Italy, where the expected number of cases without lockdown was estimated to be 669,813 compared to the observed count of 139,422, and a reduction of 26 cases (expected 43.3, observed 17.3) per 100 tests by April 8^th^, 2020. Considering that our results suggest an estimated mortality rate from COVID-19 of 10.68%, physical distancing measures may have contributed to an estimated 73,691 fewer deaths by COVID-19 in Europe alone.

On average, NPI measures showed an effect 11.05 days after the first intervention started. However, most of the countries that had a significant decrease in COVID-19 growth rates implemented NPI measures early in the start of the pandemic. This could explain why some countries may have been less impacted by NPI measures according to our trend analysis, particularly those countries that implemented lockdowns later in the time span of our analysis. We also identified countries where the lockdown did not show a significant change in the speed of the propagation of the virus, notably in France where the expected number of cases without physical distancing measures was 98,863 compared to the observed count of 90,848. This result might be linked to the lack of enforcement of the NPI measures by the government. The differences between the observed and expected number of cases across countries suggest that the impact of NPI measures on the spread of COVID-19 is dependent on the level of enforcement and intensity of the implementation of the intervention^7,15^. While countries like Italy, South Korea and Malaysia enforced strict physical distancing measures early in the pandemic, countries such as France and the United Kingdom opted for more relaxed implementation of NPI measures. Many countries are planning to start relaxing the NPI measures they currently have in place, and thus our results are potential indicators of the effectiveness of these interventions and the likely trend of the pandemic under less strict enforcement of physical distancing measures.

Despite our findings several limitations of our study are worth noting. Assumptions of our time series analysis include that the temporal trends are stable during the pre and post NPI periods^11^. In our case, this assumption is not completely accurate as the temporal trend was exponential only in the early phase of the pandemic (below 36% of total population infected^16^). This limits our analysis to short time periods after the implementation of NPI policies. Additionally, the accuracy of TPR as an approximation to COVID-19 incidence decreases if the relationship between TPR and incidence is nonlinear, which in turn would affect our interpretations regarding the impact of NPIs on COVID-19 incidence. Finally, we limited our analysis to countries with sufficient testing data to enable interpolation, and we did not consider other aspects of NPI implementations such as cultural values, level of enforcement, and frequency or magnitude of group events that could affect the propagation of the COVID-19 pandemic.

Notwithstanding these limitations, our study estimated the positive effects of NPI measures such as physical distancing orders and lockdowns in slowing the speed of the spread of the COVID-19 pandemic^3^. However, the effectiveness of NPI measures depends on the level of enforcement and intensity of the implemented interventions, and relaxed low-level enforcement of NPI measures may have a modest or even close to no impact in the course of the COVID-19 pandemic^15^. The results generated in this study could inform guidelines for the next phase of the pandemic in which intermittent physical distancing with different intervention intensity is implemented in several countries. Moreover, testing trends should be considered to assess the temporal dynamics of the pandemic. Analysis of only confirmed cases could distort the results caused by the effects of testing due to the availability of test kits, clinical guidelines, among others. Transparency in the information about testing protocols is required in order to evaluate the real spread of the pandemic and the real effects of the current NPI measures implemented.

## Data Availability

The data that support the findings of this study are openly available in the Johns Hopkins University Center for Systems Science and Engineering (https://github.com/CSSEGISandData/COVID-19)

## Acknowledgements

The authors thank the Johns Hopkins University Center for Systems Science and Engineering and the Our World in data Project for releasing the data for cases and testing for the COVID19.

## Author Contributions

Conceptualization: A.H., E.C., H.K, A.J.B., F.D.M., N.M., D.F.C.; methodology: A.H., E.C., H.K, D.F.C.; software: A.H., E.C.; validation: A.H., E.C. D.F.C.; writing—original draft preparation: A.H., D.F.C., writing—review and editing: E.C., H.K, A.J.B., F.D.M., N.M.

